# Gradient Boosting Decision Tree Algorithm for the Prediction of Postoperative Intraocular Lens Position in Cataract Surgery

**DOI:** 10.1101/2020.08.26.20181156

**Authors:** Tingyang Li, Kevin Yang, Joshua D. Stein, Nambi Nallasamy

**Affiliations:** Department of Computational Medicine and Bioinformatics, University of Michigan, Ann Arbor, MI; Kellogg Eye Center, Department of Ophthalmology and Visual Sciences, University of Michigan, Ann Arbor, MI; Center for Eye Policy and Innovation, University of Michigan, Ann Arbor, MI; Department of Health Management and Policy, University of Michigan School of Public Health, Ann Arbor, MI

## Abstract

**Purpose:** To develop a method for predicting postoperative anterior chamber depth (ACD) in cataract surgery patients based on preoperative biometry, demographics, and intraocular lens (IOL) power.

**Methods:** Patients who underwent cataract surgery and had both preoperative and postoperative biometry measurements were included. Patient demographics and IOL power were collected from the Sight Outcomes Research Collaborative (SOURCE) database. A gradient boosting decision tree model was developed to predict the postoperative ACD. The mean absolute error (MAE) and median absolute error (MedAE) were used as evaluation metrics. The performance of the proposed method was compared to five existing formulas.

**Results:** 847 patients were assigned randomly in a 4:1 ratio to a training/validation set (678 patients) and a testing set (169 patients). Using preoperative biometry and patient sex as predictors, the presented method achieved an MAE of 0.106 ± 0.098 (SD) on the testing set, and a MedAE of 0.082. MAE was significantly lower than that of the five existing methods (p < 0.01). When keratometry was excluded, our method attained an MAE of 0.123 ± 0.109, and a MedAE of 0.093. When IOL power was used as an additional predictor, our method achieved an MAE of 0.105 ± 0.091 and a MedAE of 0.080.

**Conclusions:** The presented machine learning method achieved accuracy surpassing that of previously reported methods in the prediction of postoperative ACD.

**Translational Relevance:** Increasing accuracy of postoperative ACD prediction with the presented algorithm has the potential to improve refractive outcomes in cataract surgery.

## Introduction

Estimates of postoperative intraocular lens (IOL) position after cataract surgery have been included in IOL calculations as far back as first-generation IOL formulas. Initially, postoperative IOL axial position in the eye was modeled as a constant (4mm) in anterior chamber intraocular lens (ACIOL) power calculations. In second-generation formulas, Binkhorst introduced axial length as a predictor, while third-generation formulas involved both corneal power and axial length as predictors of postoperative IOL position. By 1995, Olsen et al. introduced two additional variables, preoperative anterior chamber depth (ACD) and preoperative crystalline lens thickness as predictors for postoperative IOL position.

The importance of postoperative IOL position in IOL power calculations is due to the reliance of optical models of the eye on the distances between the optical components of the eye (the cornea and IOL) and the photoreceptors within the retina. Whether utilizing Gaussian optics or raytracing, optical models used in IOL power calculation require accurate estimates of postoperative IOL position to achieve useful results. Indeed, Norrby estimated in 2008 that estimates of IOL position were responsible for 36% of the error in IOL power predictions.^1^

While effective lens position (ELP) refers to the distance between the anterior surface of the cornea and the principal plane of the IOL resulting in the observed refraction in a given optical model, it is important to distinguish this entity from the postoperative ACD. The postoperative ACD is a measurable quantity, representing the distance between the anterior corneal surface and the anterior IOL surface along the visual axis in the postoperative eye. The relationship between ELP for a given optical model and postoperative ACD depends on the optical model itself, meaning that while postoperative ACD is a measurable quantity, ELP is only a computable quantity.

As Kriechbaum et al. pointed out in 2003, exact postoperative ACD prediction based on preoperative biometry data is, in principle, impossible because of the effect of several uncertain parameters including the shrinkage of the capsular bag.^2^ There have, however, been reports of various preoperative features that may be predictive of postoperative ACD. For example, Plat in 2017 reported correlation between measurements of axial length (AL), horizontal white to white distance (WTW), and preoperative ACD with postoperative ACD.^3^ Other approaches have added corneal power to improve postoperative ACD prediction.^4^

Methods utilizing measures from anterior segment optical coherence tomography (AS-OCT) have achieved high accuracy in postoperative ACD prediction.^5,6^ However, these approaches rely on angle-to-angle measurements that are not typically obtained in a standard cataract surgery preoperative workup. Furthermore, these measurements involve manual caliper-based measurements, introducing subjectivity and variability into the measurements while slowing the workflow of the cataract surgeon.

An ideal method for postoperative ACD estimation would utilize only data obtained from optical biometry. It would achieve high accuracy, yet have minimal loss of accuracy in the absence of reliable keratometry data. Such a method would be able to integrate into existing workflows. It could also be used for patients who had previously undergone refractive surgery, and would lend itself well to integration into existing and novel methods for IOL power calculation.

Since it is not particularly common to obtain biometry both preoperatively and postoperatively, building a dataset large enough to accurately predict postoperative ACD can be a challenge. Here, leveraging the Sight Outcomes Research Collaborative (SOURCE) database, we describe the creation of a dataset including over 800 patients with both preoperative and postoperative biometry. Furthermore, we present here the development and testing of a machine learning approach to postoperative ACD prediction.

## Methods

### Data Collection

Biometry records (including preoperative and postoperative biometry) between August 25, 2015 and June 27, 2019 were retrieved from Lenstar LS900 optical biometers (Haag-Streit USA Inc, EyeSuite software version i9.1.0.0) at University of Michigan’s Kellogg Eye Center. Institutional review board approval was obtained for the study and it was determined that informed consent was not required because of its retrospective nature and the anonymized data utilized in this study. The study was carried out in accordance with the tenets of the Declaration of Helsinki. Patient demographics and cataract surgery information (including date of surgery and implanted IOL power) were obtained via the Sight Outcomes Research Collaborative (SOURCE) Ophthalmology Data Repository, which captures electronic health record (EHR) data of all patients receiving any eye care at academic medical centers participating in this research collaborative. SOURCE captures information on patient demographics, diagnoses identified based on International Classification of Diseases (ICD) codes, procedures based on Current Procedural Terminology (CPT) codes, and structured and unstructured (free-text) data from all clinical encounters (clinic visits, operative reports, etc.). For this study, we focused on a subset of the SOURCE patients receiving care at the University of Michigan. Spherical equivalent manifest refractions from the postoperative month one visit were identified from the clinical record for all patients who underwent cataract surgery (CPT = 66984 or 66982) from the dataset. The power and model of the implanted intraocular lens for each surgery was collected as well. Only those surgeries involving the implantation of an Alcon SN60WF single-piece acrylic monofocal lens (Alcon, USA) were included in the study. Patients who had prior refractive surgeries were excluded from the dataset. Patients who had an additional surgery (e.g., endothelial keratoplasty) at the time of their cataract surgery were also excluded. Postoperative biometry records with outliers in IOL thickness were excluded to address the possibility of lens tilt affecting the assessment of ACD. The outliers were defined as records where the postoperative lens thickness fell greater than 1.96 standard deviations away from the mean thickness for a given IOL power.

### Model Development

After data collection, the raw data was reformatted so that each sample in the dataset consisted of a set of predictors and a target value that could be utilized by the machine learning model. Among the biometry records, it was possible for individual eyes to have multiple preoperative and postoperative sets of biometry measurements. In order to take advantage of these records, preoperative and postoperative biometry records of the same eye were matched in a way that accounted for all possible combinations. An eye with *x* preoperative records and *y* postoperative records had *xy* possible combinations. At the end of data preprocessing (**Figure 1**, middle panel), a dataset of 4137 samples that involved 847 distinct patients was generated and used for the development of the machine learning model. Each sample consisted of (1) preoperative biometry: axial length (AL), central corneal thickness (CCT), anterior chamber depth (ACD), crystalline lens thickness (LT), flat keratometry K1, steep keratometry K2, 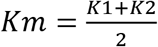, and horizontal white-to-white (WTW), (2) patient sex, (3) IOL power, and (4) postoperative ACD, where (1)-(3) were the predictors and (4) was the target variable in the machine learning model.

Corneal power is one of the most important features in both postoperative ACD prediction and postoperative refraction prediction in cataract surgery. However, corneal power measurement is unreliable in patients with prior corneal refractive surgery. In order to evaluate applicability of our method to patients with prior corneal refractive surgery, we examined how well our method performed when corneal power was not available.

We also studied the effect of IOL power in postoperative ACD prediction, because even though IOL power is directly associated with IOL thickness, which could in turn affect postoperative ACD, IOL power, to our knowledge, has not been considered in existing formulas.

In summary, we examined the performance of three classes of models where different subsets of variables were used as predictors: (1) Base, which used AL, CCT, ACD, LT, K1, K2, Km, WTW, and patient sex as predictors, (2) Base + IOL, which added IOL power to “base” as an additional feature, and (3) Base – K, which removed K1, K2, and Km from “Base”, using AL, CCT, ACD, LT, WTW, and patient sex as predictors.

LightGBM (2.2.3), which is a widely used framework for implementing the gradient boosted decision tree algorithm, was used to construct the machine learning model. During the training process, the training data were augmented through two methods (**Figure 1**, right panel) (1) IOL power augmentation, and (2) data interpolation. The purpose of using IOL power augmentation was to improve the prediction performance by incorporating the relationship between IOL power and IOL thickness into the training data. During IOL power augmentation, the implanted IOL power (*IOL_old_*) was replaced by *n_IOL_* randomly selected IOL powers, and the ground truth postoperative ACD was adjusted based on the selected IOL powers. For each distinct patient, *n_IOL_* synthetic IOL powers *(IOL_new1_,10L_new_,2*,…) between [*IOL_min_, IOL_max_*] were selected and the new postoperative ACD corresponding to each new IOL power was calculated as:

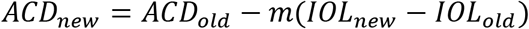

where *m* ∈ [0,1] is a constant, *IOL_min_ ≥ 6,IOL_max_ ≤* 30. The above equation assumes a simple linear relationship between the IOL power and the lens thickness: *LT_new_ − LT_old_ = m′*(*p_new_ − p_old_*), and assumes an imaginary anchor point on the visual axis within the IOL that has a fixed location independent of the IOL power. Let *LT_IOL_* be the thickness of the IOL. Two physiologically plausible assumptions about the location of IOL are that (**Figure S1**) (1) the location of the center of the IOL is fixed, meaning that the anchor point is at 0.5 * *LT_IOL_* and the distance from the cornea to the anchor point is *ACD* + 0.5 * *LT_IOL_* and (2) the location of the posterior surface of the IOL is fixed, meaning that the anchor point is at 1 * *LT_IOL_* and the distance from the cornea to the anchorage point is *ACD* + 1 * *LT_IOL_*. Here, the above assumptions were replaced by a more flexible assumption where the anchor point was at *m″ * LT_IOL_ (m″* ∈ [0,1] and *m = m′m″*). The value of *IOL_min_*, *IOL_max_*, m, and *n_I0L_* were optimized through cross-validation. In data interpolation, *k* samples were randomly picked, and the center of those *k* samples was calculated by averaging each dimension of the predictor vector *X* and the target value. Categorical variables were treated as continuous variables. The number of samples, *k*, used to create each synthetic sample and the number of samples generated*, n*, were optimized through cross-validation.

### Model Evaluation

Five repetitions of five-fold cross-validation were used to perform a grid-search for the parameters in data augmentation (*IOL_min_*, *IOL_max_*, *m*, *n_I0L_, n*, and *k*) and the hyperparameters in the machine learning model (the learning rate, number of estimators, maximum tree depth, and number of leaves). Cross-validation was also used to evaluate the performance of different subsets of features. Mean absolute error (MAE) in postoperative ACD prediction was used as the primary evaluation metric in cross-validation. The optimal models for three scenarios: (1) Base (2) Base + IOL (3) Base - K were selected based on the mean of the MAEs in the cross-validation results.

We then tested the performance of our model on a hold-out testing dataset and compared the performance of our methods with five existing formulas: Haigis, Hoffer Q, Holladay I, Olsen, and SRK/T. These five existing formulas were implemented in Python 3 based on their publications.^7–13^ The lens constants were optimized for each formula to eliminate systematic errors in refraction prediction using previously described methods.^11,14,15^ The optimized constants were: 1.655 for Haigis, 5.844 for Hoffer Q, 1.990 for Holladay I, -0.225 for Olsen, and 119.303 for SRK/T. The corresponding mean errors in refraction are listed in **Table S1**. We further compared our methods to two baseline prediction methods: (1) average postoperative ACD, which used the average postoperative ACD in the training/validation dataset as the predicted ACD for the testing set and (2) linear regression, which used AL, CCT, ACD, LT, K1, K2, Km and WTW as predictors. Data augmentation (i.e., interpolation and IOL augmentation) was not applied to the linear regression model.

To investigate the degree to which dataset size affected prediction performance, the performance of our method and linear regression were compared as random subsets of the training dataset of varying size (20%, 40%, 60%, 80%, and 100%) were utilized. The subsampling of the training dataset was applied before data interpolation to better simulate the reduction in available raw data.

During the testing and validation process, one testing/validation sample was randomly selected for each patient to ensure that performance evaluation was not biased due to the varying number of records available per patient. In addition to the MAE, the median absolute error and Pearson correlation coefficient (*r*) were also calculated for the performance comparison in the testing set. To gain insights into the relative importance of predictors in the machine learning model, we calculated the total gain (total reduction in training loss) across splits in decision trees for each predictor in the model.

### Statistical Analysis

Statistical testing was performed to investigate relationships between variables in the dataset. A chi-square test was performed to evaluate the difference in the proportion of males and females among all patients. A two-tailed Student t-test was performed to evaluate for differences in the means of biometry values between males and females. The Pearson correlation coefficients and the p-values testing the significance of correlation were calculated between the postoperative ACD and the preoperative biometry measurements. To assess the difference in cross-validation results of different methods, a Wilcoxon signed rank test was performed. The testing set results of different methods were compared based on the Friedman test followed by a post-hoc Wilcoxon signed rank test with Bonferroni correction. Statistical significance for all above tests was defined as p-value < 0.05. All statistical analysis and machine learning model construction scripts were written in Python 3.

## Results

### Dataset Characteristics

**Figure 1:**
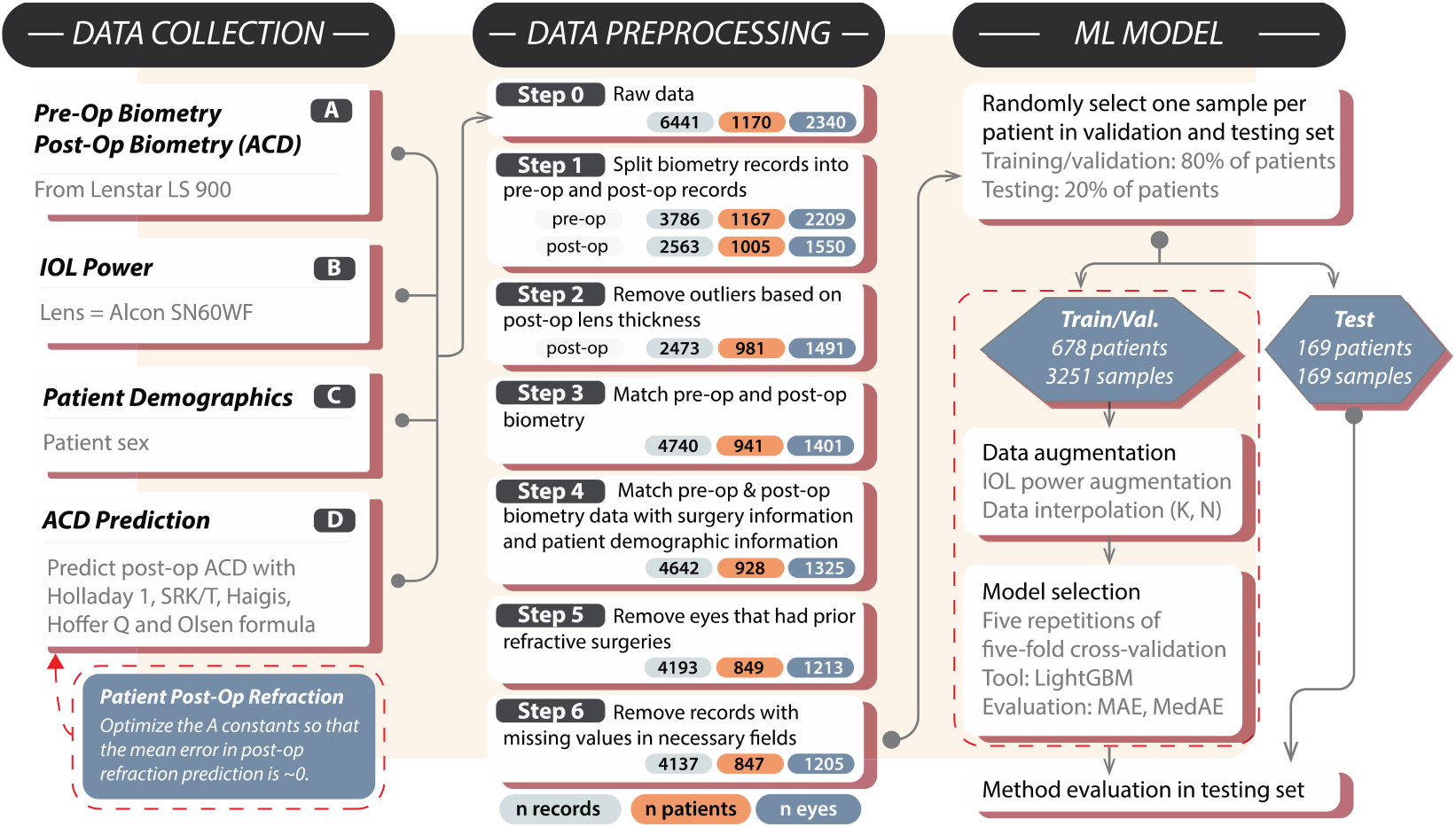
Method pipeline. In the middle panel, “n records” refers to the total number of records or samples in the whole dataset; “n patients” refers to the total number of distinct patients; “n eyes” refers to the total number of distinct eyes. In the right panel, 3251 in the training/validation set is total number of samples before selecting one sample per patient in the validation set.

**Figure 2:**
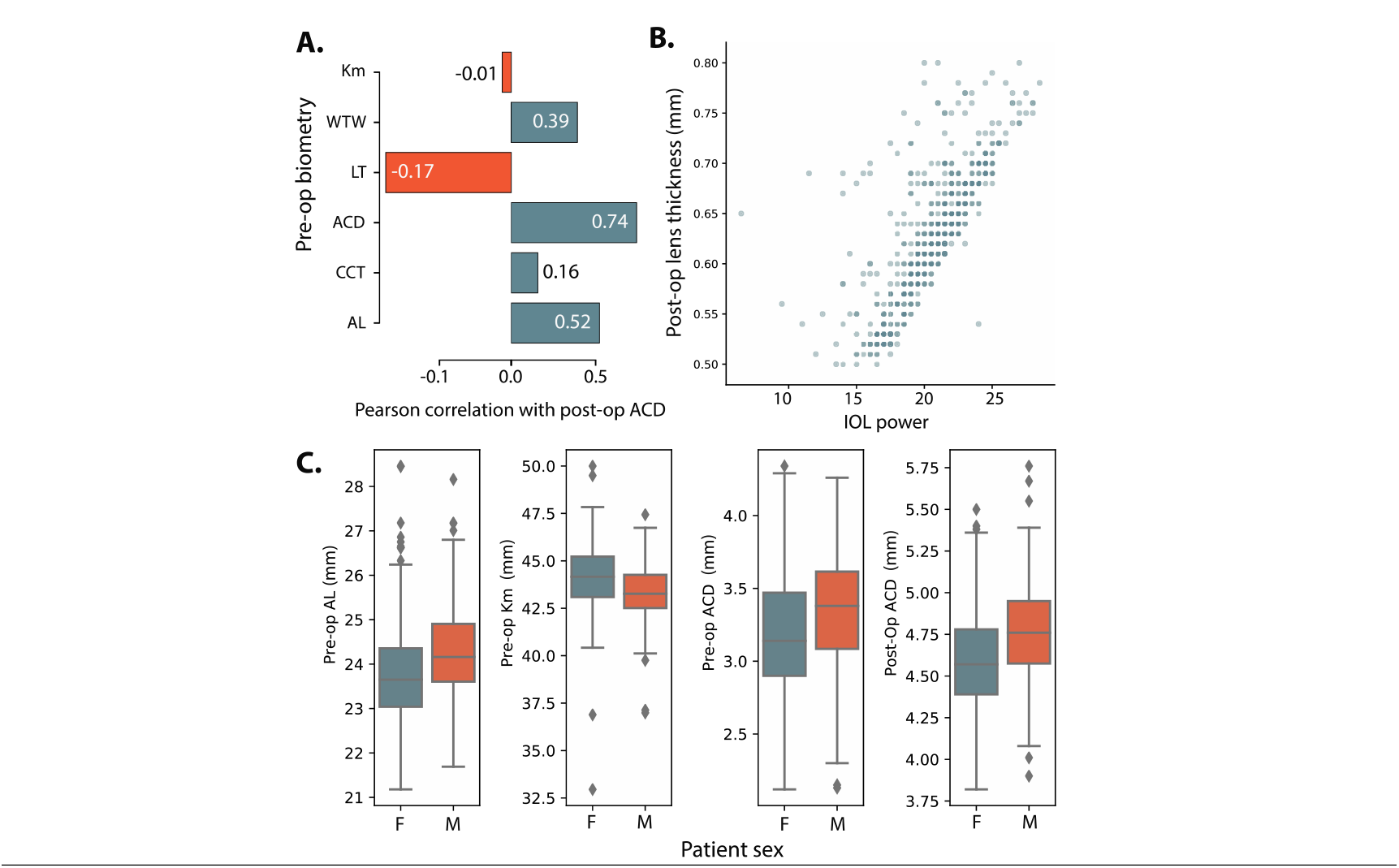
Baseline dataset characteristics. **A)** Bar graph plotting the Pearson correlation coefficient *r* between postoperative anterior chamber depth and preoperative biometry in the training/validation dataset. **B)** Scatter plot of IOL power against the postoperative lens thickness. The dots are 50% transparent. **C)** The distribution of preoperative axial length, corneal power, anterior chamber depth and postoperative anterior chamber depth in male (M) and female (F) patients. One record per patient in the training/validation set was randomly selected to generate the figures (i.e. the same set of records as the “Training/Validation Set” column in Table 1).

**Table 1:** Patient Demographics Since patients in our dataset had varied numbers of biometry exam records, we randomly selected one record/patient from the training/validation set and the testing set to calculate the summary statistics.

| Characteristic | Training/Validation Set<br>(mean ± SD) | Testing Set (mean ± SD) |
| --- | --- | --- |
| Gender | male: 283 (41.7%),<br>female: 395 (58.3%) | male: 65 (38.5%),<br>female: 104 (61.5%) |
| Age at surgery (years) | 71.08 ± 10.50 | 71.02 ± 8.96 |
| Preoperative Km (D) | 43.78 ± 1.65 | 43.94 ± 1.70 |
| Preoperative AL (mm) | 23.98 ± 1.09 | 23.79 ± 1.09 |
| Preoperative LT (mm) | 4.52 ± 0.45 | 4.53 ± 0.44 |
| Preoperative ACD (mm) | 3.25 ± 0.41 | 3.24 ± 0.42 |
| Postoperative ACD (mm) | 4.66 ± 0.30 | 4.64 ± 0.30 |

In total, our dataset included the preoperative and postoperative biometry measurements and surgical records of 1205 eyes from 847 patients (**Figure 1**). These patients were split into training/validation and testing sets. The distributions of the biometry measurements were similar in the two sets (**Table 1**). There were significantly more females than males (chi-square test, p < 0.01). The postoperative anterior chamber depth (ACD) was positively correlated with preoperative AL, ACD, WTW, and CCT (p < 0.01 for each), and negatively correlated with preoperative LT and WTW (p < 0.01 for each). Postoperative ACD was not significantly correlated with preoperative Km (p = 0.74) (**Figure 2A**). **Figure 2B** shows the distribution of the power of the implanted IOL and the postoperative lens thickness (*r* = 0.75, p < 0.01). The scatter plot indicates a linear relationship between the IOL power and postoperative IOL thickness. The distributions of biometry measurements in male and female patients are shown in **Figure 2C**. The preoperative AL, preoperative ACD, and postoperative ACD in male patients were longer than those in female patients (p < 0.01 for each). Km in females was greater than that in males (p < 0.01).

### Model Performance

**Table 2:** Prediction Performance on the Testing Set MAE = Mean Absolute Error. MedAE = Median Absolute Error.

| Index | Method | MAE in mm ± SD | MedAE in mm (interquartile range) | $R^2$ |
| --- | --- | --- | --- | --- |
| 1 | Base = biometry + patient sex | 0.106 ± 0.098 | 0.082 (0.119) | 0.777 |
| 2 | Base + IOL | 0.105 ± 0.091 | 0.080 (0.114) | 0.781 |
| 3 | Base – K | 0.123 ± 0.109 | 0.093 (0.124) | 0.711 |
| 4 | Haigis | 0.680 ± 0.172 | 0.681 (0.206) | 0.681 |
| 5 | Hoffer Q | 1.228 ± 0.251 | 1.219 (0.318) | 0.407 |
| 6 | Holladay I | 0.743 ± 0.283 | 0.744 (0.403) | 0.405 |
| 7 | Olsen | 1.200 ± 0.172 | 1.199 (0.206) | 0.681 |
| 8 | SRK/T | 1.205 ± 0.328 | 1.183 (0.256) | 0.317 |
| 9 | Average postoperative ACD | 0.231 ± 0.195 | 0.192 (0.000) | / |
| 10 | Linear regression | 0.116 ± 0.099 | 0.089 (0.120) | 0.746 |

As stated above, different subsets of features were tested to examine the performance of our machine learning model when (1) corneal power was not available and (2) IOL power was considered. **Figure S2** shows the cross-validation results of the alternative models with optimized parameters. The cross-validation results (i.e., the average MAE) of each alternative model were as follows: 0.121 mm for Base = biometry + patient sex, 0.120 mm for Base + IOL, 0.131 mm for Base — K. The addition of IOL power improved prediction performance, while IOL-based augmentation, which simulated the linear relationship between an IOL’s power and its thickness further improved prediction performance, beyond the addition of IOL power alone. Base – K performed significantly worse compared to Base and Base + IOL (p < 0.01), as expected. For comparison purposes, we recalculated the cross-validation results using median absolute error as the evaluation metric. The results were as follows: 0.100 mm for Base, 0.097 mm for Base + IOL, and 0.108 mm for Base – K. The prediction performance was consistent with the results obtained with MAE.

The performance of the three models on the unseen testing dataset is presented in **Table 2**. The Friedman test for difference in MAE among the methods in Table 2 was significant (p < 0.01). The Base predictors, which included preoperative biometry and patient sex, achieved an MAE of 0.106 mm. Adding the IOL improved the prediction performance in the test set (MAE = 0.105 mm). Base and Base + IOL significantly outperformed Haigis, Hoffer Q, Holladay I, Olsen, SRK/T, and mean postoperative ACD, based on the post-hoc Wilcoxon signed rank test with Bonferroni correction (p < 0.01). When the corneal power was not included (Base − K), which simulates the scenario when the measured corneal power is not reliable, our method maintained good performance, with an MAE = 0.123 mm. The performance of Base - K still significantly outperformed the existing 5 formulas (p < 0.01).

**Figure 3:**
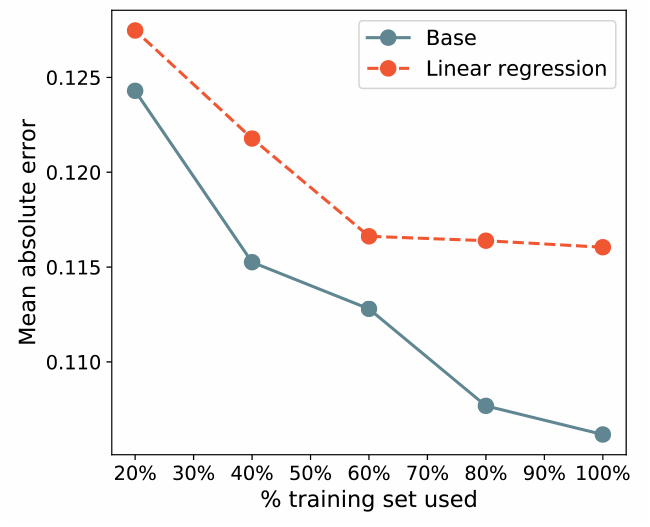
Testing set performance (MAE of postoperative ACD prediction in mm) of the linear regression method (dashed line) and our Base method (Base = biometry + patient sex) (solid line). MAE = mean absolute error, ACD = anterior chamber depth.

The performance of our methods and the linear regression method on training datasets of varying size is shown in **Figure 3**. The result demonstrates that the performance attained by our Base method on the testing set continued to improve as the dataset grew to 100% of the available data. On the contrary, the improvement of the linear regression method plateaued at around 60% of the available data. The above result indicates that the large set of paired preoperative and postoperative biometry provided a significant benefit to our machine learning model and that its performance may continue to improve as more data becomes available.

**Figure 4:**
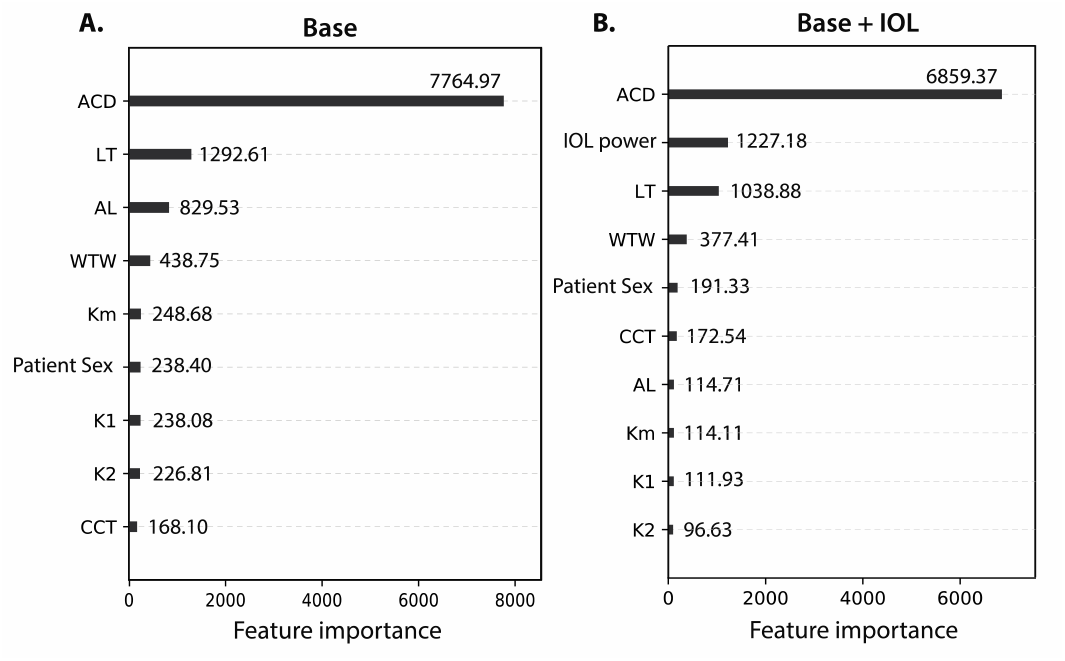
Feature importance in Base model and Base + IOL model, measured by total gain across splits.

Feature importance in the Base model and Base model with IOL power is shown in **Figure 4**. ACD, LT, and WTW ranked highly in both models. IOL power achieved a high importance score when it was added into the model (**Figure 4B**).

## Discussion

We have presented here a machine learning approach to predicting postoperative ACD in cataract surgery using standard preoperative optical biometry measurements.

In order to develop this method, we built, to our knowledge, the largest optical biometry dataset involving both preoperative and postoperative measurements reported to date. We found, through sampling of subsets of this dataset of varying size, that the performance of our machine learning method for predicting postoperative ACD improved substantially as the dataset size grew, while the performance of linear regression plateaued at 60% of the available data. This finding underscores the potential of our machine learning method to continue to improve as the dataset, derived from the SOURCE data repository, continues to grow.

We found that even a linear regression approach to modeling postoperative ACD achieved performance better than that of previously reported methods (including Haigis, Hoffer Q, Holladay I, Olsen, and SRK/T) both in terms of the mean absolute error and the Pearson correlation (*r*). The performance of linear regression on our biometry dataset also exceeded that of previously reported AS-OCT methods by *R*^2^ value.^5^ Since lens constants were optimized individually prior to computing the predictions of each of the aforementioned formulas, the high performance of linear regression relative to existing methods was likely due to the size of the dataset of available, as well as the use of optical biometry to directly measure postoperative IOL position, as opposed to ultrasound biometry or ELP calculations. The existing formulas considered here use a thin lens assumption, wherein the intermediate value referred to as the ACD does not represent the position of either surface of the IOL, but rather the location of the principal plane.^4^ Therefore, the estimated ACD terms in these formulas can more accurately be described as providing information about the ELP within the optical models employed by those IOL power calculation formulas. As such, they are not ideal for prediction of the true postoperative anatomy of the eye of a cataract surgery patient.

By employing a gradient boosting machine learning algorithm, we were able to significantly improve ACD prediction performance beyond that of linear regression and existing ACD prediction formulas. Our method also outperformed methods employing more involved measurement techniques such as AS-OCT.^6,16^ Evaluation of feature importance demonstrated that preoperative ACD was the most important input parameter, followed by crystalline lens thickness (LT), axial length (AL), and horizontal white to white (WTW), respectively. Inclusion of patient sex, which is not typically utilized in methods of postoperative ACD prediction, in the model was found to improve performance (**Figure S2**). This finding was consistent with prior studies of patient biometry reporting consistent differences in ocular shape between male and female patients, with female corneal powers measuring greater and axial lengths measuring shorter than those of males on average.^17,18^

In order to enhance performance of our gradient boosting machine learning approach, we employed two techniques for data augmentation. One of these methods involved modeling IOL thickness based on IOL power to account for potential variations in postoperative ACD due to the thickness of the IOL utilized. Both of the data augmentation methods presented here resulted in improvements cross-validation performance (**Figure S2**). The performance enhancement seen with the IOL thickness adjustment method indicates that IOL thickness is indeed relevant to postoperative ACD. It further indicates that customized IOL thickness modeling should be included in methods for postoperative ACD prediction depending on the model of IOL intended for use in the patient’s preoperative plan.

Due to challenges in accurately assessing corneal power in post-refractive surgery patients, we investigated a keratometry-independent (K-independent) approach to prediction of postoperative ACD as well. Our method outperformed a previously-reported method for K-independent prediction of ACD,^19^ and may be applicable in new methods for IOL power calculation in patients with prior refractive surgery.

The limitations of our study include the use of a retrospective, rather than prospective, dataset. It was not possible to compare our method for ACD prediction to those of the Barrett Universal II or Holladay 2 formulas, as the ACD predictions of these formulas are not publicly available. An additional limitation of our study is that while a hold-out testing set was used, it was comprised of data obtained at the same institution. Building a separate testing set external to our institution would provide additional validation of our method, and will be a future direction of this work.

The method presented here for prediction of postoperative ACD has the potential to be integrated into methods for IOL power calculation. Both geometrical optics and ray-tracing methods for IOL power calculation rely on some form of prediction of postoperative IOL position, and could benefit from the accuracy of the approach presented here. Since feature engineering is an important part of optimizing machine learning methods, and postoperative ACD is known to be a useful predictor in traditional methods of IOL power calculation, it is possible that postoperative ACD may be a useful feature for machine learning approaches to IOL power calculation as well.

In summary, the machine learning method presented here for predicting postoperative ACD in cataract surgery has the potential for integration into novel methods for IOL power calculation, both in standard and post-refractive surgery cases.

## Data Availability

The raw data used in this study is not publicly available.

